# Anti-Spike protein assays to determine post-vaccination antibody levels: a head-to-head comparison of five quantitative assays

**DOI:** 10.1101/2021.03.05.21252977

**Authors:** Thomas Perkmann, Nicole Perkmann-Nagele, Thomas Koller, Patrick Mucher, Astrid Radakovics, Rodrig Marculescu, Michael Wolzt, Oswald F. Wagner, Christoph J. Binder, Helmuth Haslacher

## Abstract

**Background:** Reliable quantification of the antibody response to SARS-CoV-2 vaccination is highly relevant for identifying possible vaccine failure and estimating the time of protection. Therefore, we aimed to evaluate the performance of five different Anti-SARS-CoV-2 antibody assays regarding the quantification of anti-spike (S) antibodies induced after a single dose of BNT162b2.

**Methods:** Sera of n=69 SARS-CoV-2 naïve individuals 21±1 days after vaccination with BNT162b2 (Pfizer/BioNTech) were tested using the following quantitative SARS-CoV-2 antibody assays: Roche S total antibody, DiaSorin trimeric spike IgG, DiaSorin S1/S2 IgG, Abbott II IgG, and Serion/Virion IgG. Test agreement was assessed by Passing-Bablok regression. Results were further compared to the percent inhibition calculated from a surrogate virus neutralization test (sVNT) by correlation and ROC (receiver-operating-characteristics) analysis.

**Results:** Individual values were distributed over several orders of magnitude for all assays evaluated. Although the assays were in good overall agreement (ρ=0.80-0.94), Passing-Bablok regression revealed systematic and proportional differences, which could not be eliminated by converting the results to BAU/mL as suggested by the manufacturers. 7 (10%) individuals had a negative sVNT results (i.e. <30% inhibition). These samples were reliably identified by most assays and yielded low binding antibody levels (ROC-AUCs 0.84-0.93).

**Conclusions:** Although all assays evaluated showed good correlation, readings from different assays were not interchangeable, even when converted to BAU/mL using the WHO international standard for SARS-CoV-2 immunoglobulin. This highlights the need for further standardization of SARS-CoV-2 serology.

## Background

SARS-CoV-2 antibody testing played and still plays an essential role in the management of the COVID-19 pandemic *(1)*. Detection of specific antibodies following SARS-CoV-2 infection is important at both the individual and population levels to identify those at risk of infection *(2)*. However, now in the early vaccination era of the COVID-19 pandemic, another essential role of SARS-CoV-2 serology is added: the determination of specific antibodies after active immunization *(3, 4)*. Although this new role has not yet been finally defined, it is already clear that antibody assays will be needed for both vaccine development and approval process and the follow-up of vaccinated individuals. Licensing studies on a scale such as Pfizer/BioNtech and Moderna vaccines will not be feasible for all vaccine candidates and the companies behind them *(5)*. For this reason, correlates of vaccine-induced protection will have to replace, at least in part, the measurement of clinical outcomes as the sole measure. For reasons of simplicity, relatively good standardizability, and broad applicability, SARS-CoV-2 antibody tests have the best chance of becoming such substitute endpoints *(6)*.

The first SARS-CoV-2 antibody testing systems were designed to distinguish individuals with prior COVID-19 infection from those who were still naive to this new virus *(7)*. Therefore, these immunoassays were usually developed as qualitative rather than quantitative tests and were designed by the manufacturer to achieve the highest possible specificity and high sensitivity. High specificity was indispensable, especially at the beginning of the pandemic, because the extremely low seroprevalence rates led to many false positives and low positive predictive values even with tests having a specificity of 99% *(8)*. In contrast, the sensitivity of SARS-CoV-2 testing was often reduced to assure the high specificities needed for these assays *(9)*. The lower antibody levels further aggravated suboptimal sensitivities in mild/asymptomatic infections and during the pandemic by the natural decline in antibody levels *(10-15)*.

Various antigens have been used for this purpose, but essentially two types can be distinguished: nucleocapsid- and spike protein-based assays *(16)*. Antibodies directed against SARS-CoV-2 specific nucleocapsid (NC) antigens are induced early and strongly in most infected individuals due to the virus nucleocapsid’s typical strong immunogenicity *(17)*. Furthermore, a very high specificity can be achieved by targeted modification of the nucleocapsid antigen so that no cross-reactivity is observed even with closely related viruses. The discriminatory properties of such nucleocapsid-based antibody assays can therefore be excellent *(18, 19)*. The physiological significance of these antibodies, on the other hand, is unclear, and these surrogate markers for a previous infection are unlikely to be functionally relevant to confer protection or immunity. The antibodies that react with the spike protein (S), however, act differently. At least a proportion of these S-binding antibodies are likely to have the function of neutralizing antibodies *(20)*. Thus, it is not surprising that numerous studies have shown a correlation between spike protein binding assays and various forms of functional virus neutralization assays *(21-26)*.

In the context of SARS-CoV-2 vaccines, it is precisely these neutralizing antibodies that are of paramount importance. The primary goal of active immunization is to induce many SARS-CoV-2-specific neutralizing antibodies that ideally prevent the pathogen’s entry and thus infection or stop the systemic spread to prevent disease *(27)*. The functional virus neutralization assays are not feasible everywhere: assays with live viruses require biosafety level 3, but variants such as pseudotyped neutralization assays are also labor-intensive and cannot be performed at high throughput *(28-30)*. Classical antibody assays, which measure the reactivity of antibodies in serum/plasma with defined antigens, can be performed very rapidly and in high throughput, in contrast to neutralization tests.

Thus, anti-spike protein assays will play an important role in vaccine development, licensing, and efficacy monitoring in the future. However, these test systems must be able to reliably quantitate SARS-CoV-2 specific antibody levels, be comparable to each other, and have good to excellent agreement with the presence of neutralizing antibodies. The comparability of antibody assays is expected to be improved by the recent introduction of a first WHO International Standard for anti-SARS-CoV-2 immunoglobulin (NIBSC code 20/136) with reference to neutralizing antibodies.

In the present work, we aim to go a step further and characterize the vaccination response after the first administration of the Pfizer/BioNTech btn162b2 vaccine using five commercial quantitative anti-spike protein antibody assays (4 of them with manufacturer’s correction factor for the WHO standard) in a head-to-head comparison.

## Methods

### Study design and participants

This prospective observational study includes sera taken from 69 individuals without a previous SARS-CoV-2 infection taken 21±1 days (mean±standard deviation) after the first dose of the Pfizer/BioNTech BNT162b2 vaccine. Further inclusion criteria were an age >18 years, whereas an insufficient amount of serum would have led to exclusion from the study. The study protocol was reviewed and approved by the Ethics Committee of the Medical University of Vienna (EK1066/2021). All participants provided written informed consent to donate blood for the evaluation of diagnostic test systems (EK404/2012).

### Laboratory procedures

Serum was obtained and stored at 2 – 10°C for <7 days within the MedUni Wien Biobank, a centralized facility for the preparation and storage of biomaterial with certified quality management (ISO 9001:2015)*(31)*. All analytical procedures were performed at the Department for Laboratory Medicine, Medical University of Vienna. The following CE-marked binding assays were applied:

The Roche Elecsys® Anti-SARS-CoV-2 S (Roche S tAb) is an electrochemiluminescence sandwich immunoassay (ECLIA) and detects total antibodies directed against the receptor-binding domain (RBD) of the viral spike (S)-protein and was measured on cobas® e801 modular analyzers (Roche Diagnostics, Rotkreuz, Switzerland). The quantification range is between 0.4 and 2500.0 U/mL. The manufacturer states intra- and interassay precision between 1 and 3%, a clinical specificity of 99.98% (99.91 – 100), and a cumulative sensitivity ≥14 days after the first positive PCR of 98.8% (98.1 – 99.3) if 0.8 U/mL is used as a cut-off.

The Abbott SARS-CoV-2 IgG II Quant-test (Abbott S IgG) is a chemiluminescence microparticle immunoassay (CMIA). It quantifies IgG-type antibodies against the RBD of the viral S-protein on an Abbott ARCHITECT platform (Abbott, Abbott Park, USA) between 21.0 and 40,000.0 AU/mL. Intra- and interassay precision ranges between 3 and 5%. According to the manufacturer, clinical specificity is 99.55% (99.15 – 99.76), and clinical sensitivity is 98.81% (93.56 – 99.94) ≥15 days after the first positive PCR at a cut-off of ≥50 AU/mL.

The DiaSorin LIAISON SARS-CoV-2 TrimericS IgG (DiaSorin TriS IgG) chemiluminescence immunoassay (CLIA) quantifies IgG antibodies against a trimeric S-protein antigen on a DiaSorin LIAISON (DiaSorin, Stillwater, USA). The quantification range is between 1.63 and 800 AU/mL. Intra- and interassay precision ranges between 0 and 5%. According to the manufacturer, clinical specificity is 99.5% (99.0 – 99.7), and clinical sensitivity ≥15 days after the first positive PCR is 98.7% (94.5 – 99.6) at a cut-off of ≥13 AU/mL.

The DiaSorin LIAISON SARS-CoV-2 S1/2 CLIA (DiaSorin S1/2 IgG) detects IgG antibodies against an S1/S2 combination antigen on a DiaSorin LIAISON (DiaSorin, Stillwater, USA). The quantification range is between 3.8 and 400.0 AU/mL. Intra- and interassay precision ranges between 0 and 4% and, according to the manufacturer, specificity among blood donors is 98.5% (97.5 99.2) and sensitivity is 97.4% >15 days after diagnosis at a cut-off of >15 AU/mL, whereby results between 12.0 and 15.0 AU/mL are considered borderline.

The Virion\Serion ELISA (enzyme-linked immunosorbent assay) agile SARS-CoV-2 IgG (Serion IgG) (Institut Virion-Serion, Wuerzburg, Germany) was analyzed on a FilterMax F5 Multiplate Reader (Molecular Devices, San José, USA) and quantifies IgG antibodies against total S-protein between 3 and 250 U/mL. Intra- and interassay precision ranges between 1 and 4%. According to the manufacturer, specificity is 99.2%, and sensitivity is 96.2% at a cut-off of 15 U/mL, with values between 10 and 15 U/mL being considered borderline results.

If applicable, binding antibody units per milliliter (BAU/mL), which are traceable to the WHO International Standard for anti-SARS-CoV-2 immunoglobulin, were calculated by applying the following conversion factors, as suggested by the manufacturers: Roche 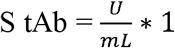, Abbott S 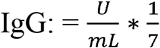, DiaSorin TriS 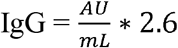, Serion 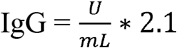.

We excluded prior SARS-CoV-2 infection by using the Roche Elecsys® SARS-CoV-2 ECLIA on the cobas® e801 analyzer (Roche), which detects total antibodies to the viral nucleocapsid antigen. These antibodies are not induced by vaccination with bnt162b2. This assay yields high diagnostic sensitivity (90%) and specificity (99.7%) for infections that occurred at least 14 days before blood withdrawal *(7)*. As suggested by the manufacturer, results >1.000 COI were considered positive.

Neutralizing capacity was estimated by performing a surrogate virus neutralization test (sVNT) (GenScript, Piscataway, USA). The assay was read on a Filtermax F5 multimodal plate reader. According to the manufacturer, it shows excellent positive (100% [87.1 – 100.0]) and negative percent agreement (100.0% [95.8 – 100.0]) with conventional plaque-reduction neutralization tests (PRNT_50_ and PRNT_90_). Results ≥30% are considered positive.

### Statistical analysis

Continuous data are given as a median and interquartile range, categorical data as counts and percentages. Passing-Bablok regressions, Cohen’s Kappa (linear weights) and Spearman rank correlations evaluated the agreement between binding assays. The relationship between binding assays and results from the sVNT was described by quadratic curve fitting. The predictive value of binding assays regarding positivity in the sVNT was assessed by interpreting and comparing (according to DeLong) the areas under the curve (AUC) from receiver-operating-characteristics (ROC)-curves. Statistical significance was assumed if *P* values were below 0.05. All analyses were performed using MedCalc 19.6 (MedCalc, Ostend, Belgium), and graphs were drawn using GraphPad 9 (GraphPad, La Jolla, USA).

## Results

### Measurement ranges differ between binding assays

29 female (42%) and 40 male (58%) participants with a median age of 42 years (29 – 51) were included. Results from the five different antibody binding assays are presented in Table 1 and Figure 1. The Abbott S IgG assay showed the highest values with a median of 1097.1 AU/mL (580.1 – 1959.5) and the broadest range (1.4 – 8281.0). In contrast, the DiaSorin S1/2 IgG CLIA yielded the lowest values (63.7 AU/mL [47.8 – 87.5]), and the levels ranged from below the limit of quantification (<3.8%, 1 sample) to 148.0. Two assays, the DiaSorin Tris IgG (195.0 AU/mL [99.0 – 337.3]) and the Serion IgG (50 U/mL [30 – 89]) returned a result above the measuring range for the same donor (>800 AU/mL and >250 U/mL). However, both tests used their full available range (lowest values: 1.8 AU/mL and <3 U/mL). Roche S tAb ECLIA results came in between the other test systems (79.6 U/mL [24.7 – 142.3]), ranging from 0.4 to 508.0.

**Tbl 1:**
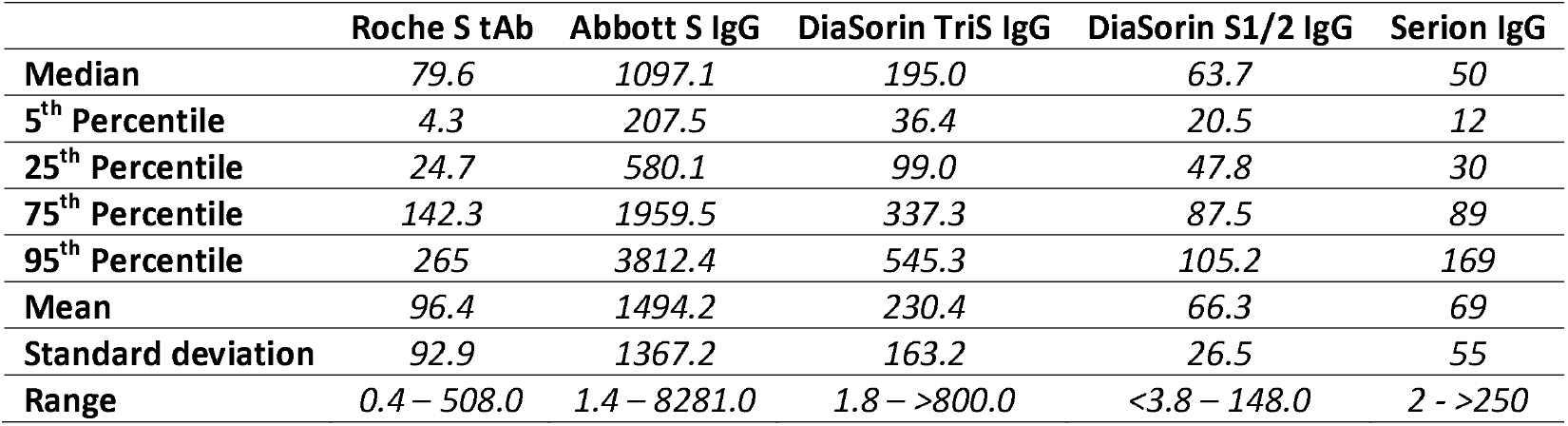
Dimensions of position and spread for 5 S-protein based SARS-CoV-2 antibody assays calculated from N=69 samples taken 21±1 days after the first shot of bnt162b2.

|  | Roche S tAb | Abbott S IgG | DiaSorin TriS IgG | DiaSorin S1/2 IgG | Serion IgG |
| --- | --- | --- | --- | --- | --- |
| **Median** | 79.6 | 1097.1 | 195.0 | 63.7 | 50 |
| **5<sup>th</sup> Percentile** | 4.3 | 207.5 | 36.4 | 20.5 | 12 |
| **25<sup>th</sup> Percentile** | 24.7 | 580.1 | 99.0 | 47.8 | 30 |
| **75<sup>th</sup> Percentile** | 142.3 | 1959.5 | 337.3 | 87.5 | 89 |
| **95<sup>th</sup> Percentile** | 265 | 3812.4 | 545.3 | 105.2 | 169 |
| **Mean** | 96.4 | 1494.2 | 230.4 | 66.3 | 69 |
| **Standard deviation** | 92.9 | 1367.2 | 163.2 | 26.5 | 55 |
| **Range** | 0.4 – 508.0 | 1.4 – 8281.0 | 1.8 – >800.0 | <3.8 – 148.0 | 2 - >250 |

**Fig 1:**
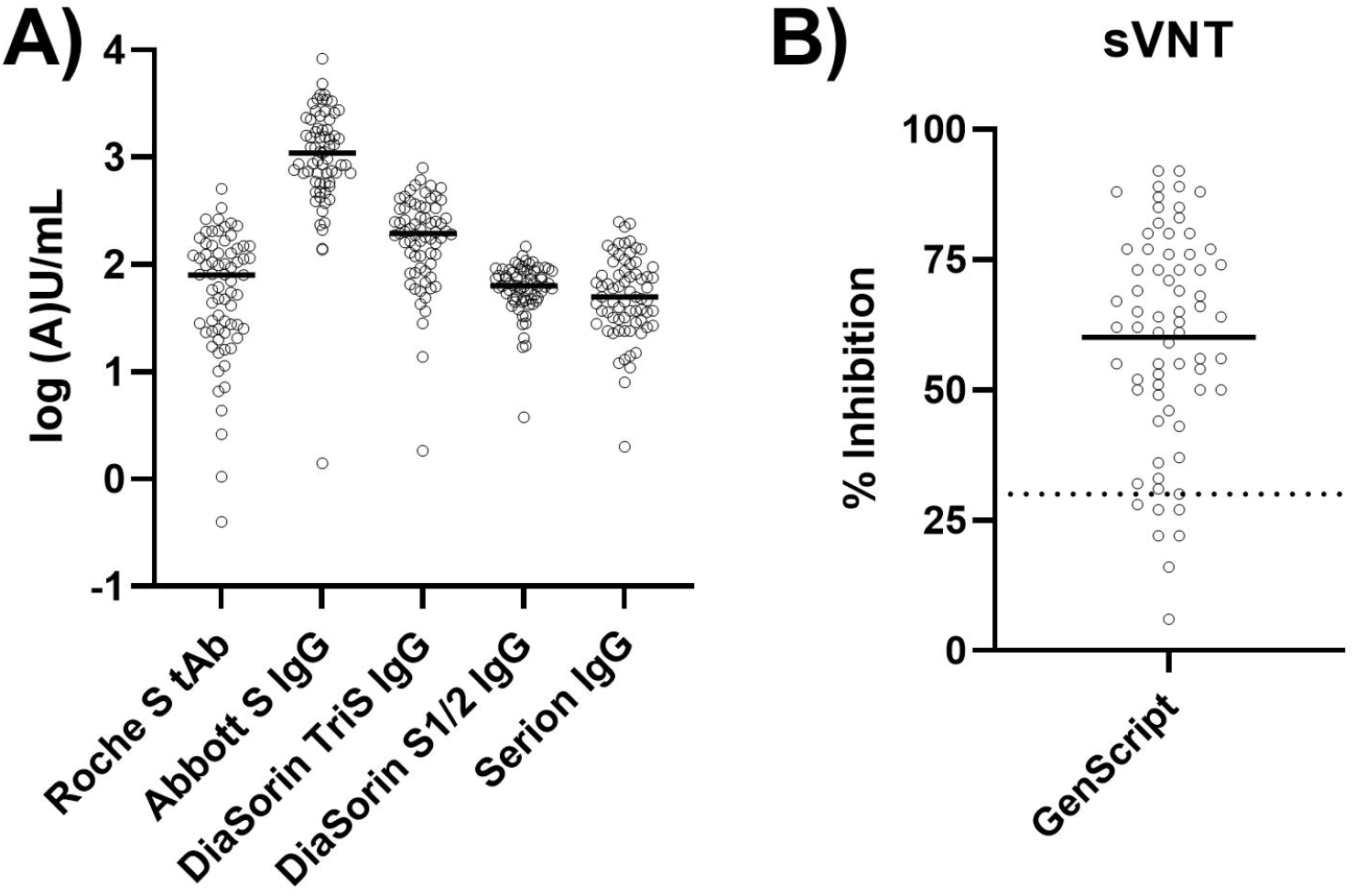
Results from binding assays **(A)** and a surrogate virus neutralization test, sVNT **(B)**. Solid lines mark the median. The dotted line in (B) marks the manufacturer’s threshold for positivity (30%).

The measured values indicate that the numerical results are strongly dependent on the test system used. In the next step, we aimed to evaluate the overall agreement between the test systems.

### Agreement between results from different binding assays

Results from the Roche S tAb assay correlated well with those of the other binding assays (Abbott S IgG ρ=0.88, DiaSorin TriS IgG ρ=0.83, DiaSorin S1/2 IgG ρ=0.80, Serion IgG ρ=0.82). However, Passing-Bablok regression revealed relevant systematic and proportional differences: Abbott S IgG = 82.5 + 15.54*X, DiaSorin TriS IgG = 33.4 + 2.18*X, DiaSorin S1/2 IgG = 39.6 + 0.32*X, Serion IgG = 12.3 + 0.65*X.

The Abbott S IgG assay correlated at ρ=0.90 with the remaining three test systems (DiaSorin TriS IgG and S1/2 IgG, Serion IgG). In Passing-Bablok regression, all systematic and proportional errors were statistically significant: DiaSorin TriS IgG = 24.5 + 0.13*X, DiaSorin S1/2 IgG = 34.5 + 0.02*X, Serion IgG = 6.2 + 0.04*X.

The DiaSorin TriS IgG assay showed an excellent correlation with the remaining two tests (DiaSorin S1/2 IgG ρ=0.91, Serion IgG 0.94). In the Passing-Bablok regression, nevertheless, marked deviations became apparent: DiaSorin S1/2 IgG = 30.5 + 0.16*X, Serion IgG = −0.0 + 0.31*X.

Finally, the DiaSorin S1/2 IgG and the Serion IgG correlated at ρ=0.91, and the Passing-Bablok regression equation was Serion IgG = −50.9 + 1.78*X. All described relationships, as well as related residual plots, are presented in Figure 2.

**Fig 2:**
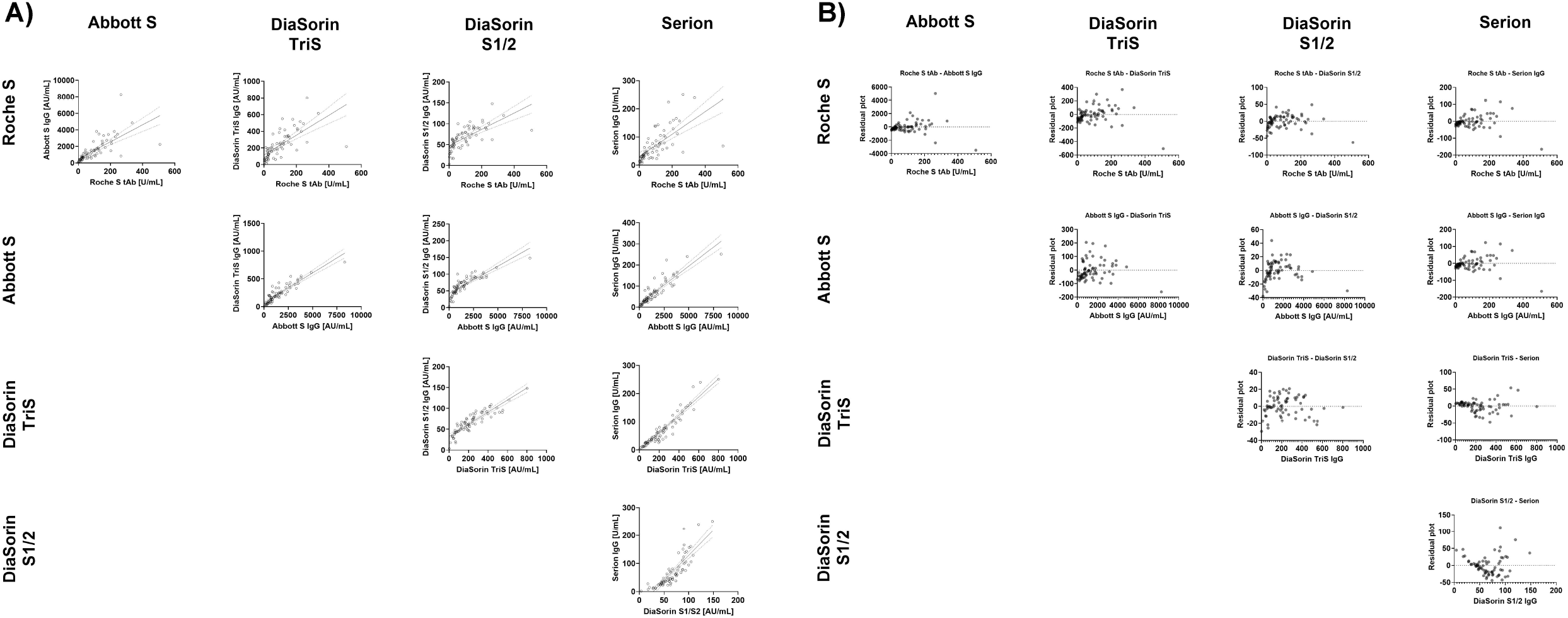
Comparison of binding assays by linear regression (dotted lines indicate the 95% confidence interval) **(A)** and residual plots **(B)**.

Furthermore, we assessed whether the classification of results into tertiles (0 – 33.3%, 33.4 – 66.7%, 66.8 – 100%) was comparable, e.g., whether a sample yielding a result in the lowest tertile of test A was also in the lowest tertile of test B. Cohen’s kappa was between 0.60 and 0.80, indicating a good agreement, for all but for one of the ten test combinations (Roche S tAb/Serion, kappa = 0.59, see Table 2).

**Tbl 2:** Kappa (±95% confidence interval) for ten different test combinations regarding the classifications of samples into tertiles.

|  | Abbott S IgG | DiaSorin TriS IgG | DiaSorin S1/2 IgG | Serion IgG |
| --- | --- | --- | --- | --- |
| **Roche S tAb** | 0.72±0.06 | 0.69±0.06 | 0.63±0.07 | 0.59±0.07 |
| **Abbott S IgG** |  | 0.64±0.07 | 0.74±0.06 | 0.64±0.07 |
| **DiaSorin TriS IgG** |  |  | 0.74±0.06 | 0.74±0.06 |
| **DiaSorin S1/2 IgG** |  |  |  | 0.80±0.05 |

In conclusion, the results of the investigated test systems correlate well but are not necessarily interchangeable. Several manufacturers provided conversion factors related to the WHO International Standard for SARS-CoV-2 immunoglobulin, as described in the methods section. Next, we wanted to clarify whether comparing converted BAU/mL instead of arbitrary values facilitates comparability.

### Associations between standardized binding assay results

Binding antibody units per milliliter (BAU/mL) were calculated for the Abbott S IgG, the DiaSorin TriS IgG, and the Serion IgG, according to the recently proposed conversion factors. Results from the Roche S tAb ECLIA did not require conversion as indicated by the manufacturer.

As shown in Figure 3, the BAU/mL recalculation did not solve the problem of high proportional errors. The least proportional error could be observed for the relationship between Roche S tAb and Serion IgG. However, the same combination was characterized by comparatively high variability (ρ=0.82).

**Fig 3:**
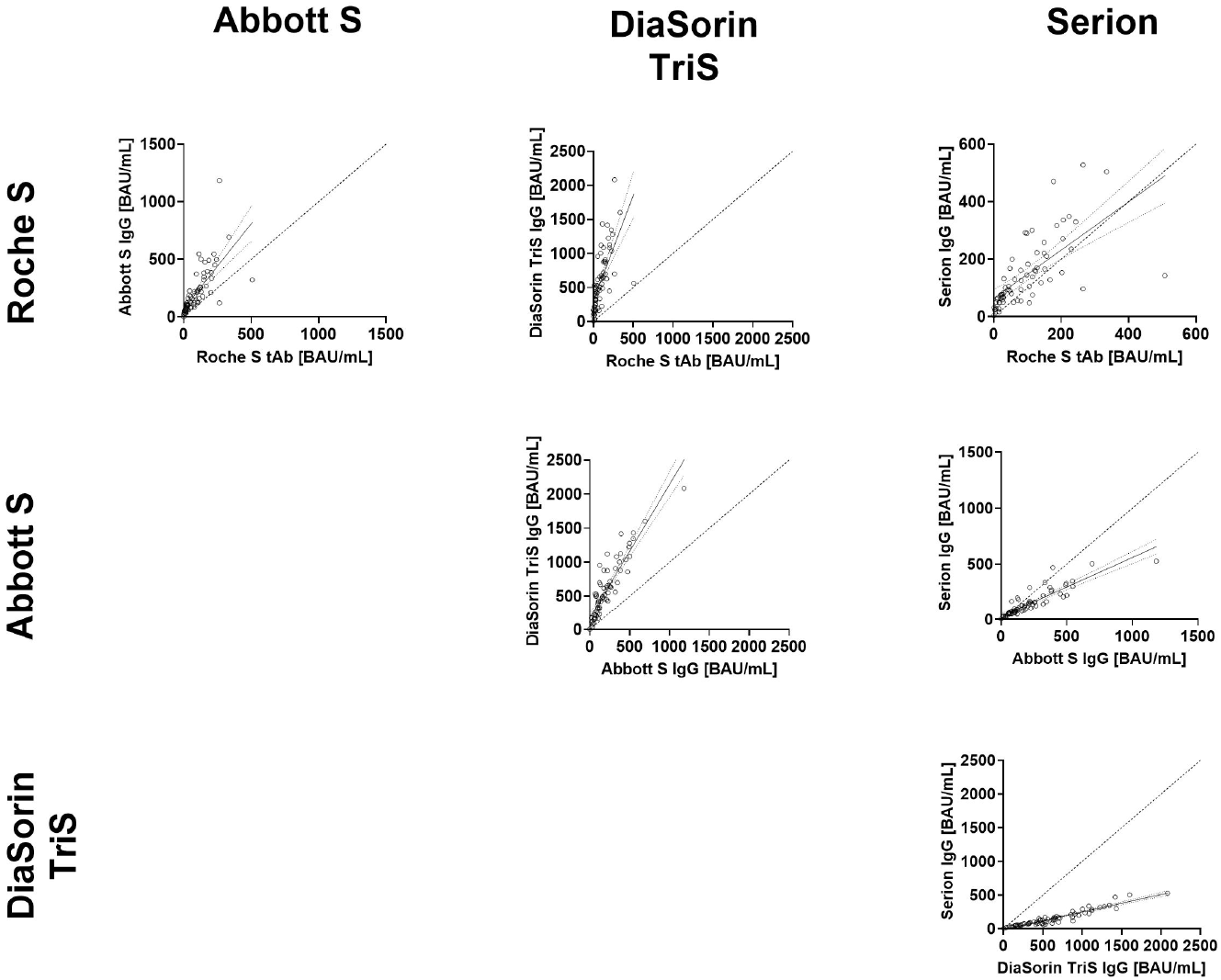
Relationships between binding assay results and percent inhibition assayed using a surrogate virus neutralizaltion test (threshold for positivity: 30%, dotted vertical lines). Presented are quadratic regression lines and their 95% confidence intervals.

### Correlation of binding assay results with a surrogate neutralization assay

In a final step, the binding assays’ results were compared to percent inhibition of a surrogate virus neutralization assay (sVNT). In the sVNT, the tested samples yielded median values of 63% (50 – 76), ranging from 6 to 92%. Figure 4a illustrates that all binding assays except the DiaSorin S1/2 IgG showed a quadratic relationship with the sVNT. The binding assays also differentiated those values clustered in the upper range of the sVNT. However, for the DiaSorin S1/2, the quadratic curve approached a straight line, indicating a mostly linear relationship between this binding assay and the sVNT within the observed range.

**Fig 4:**
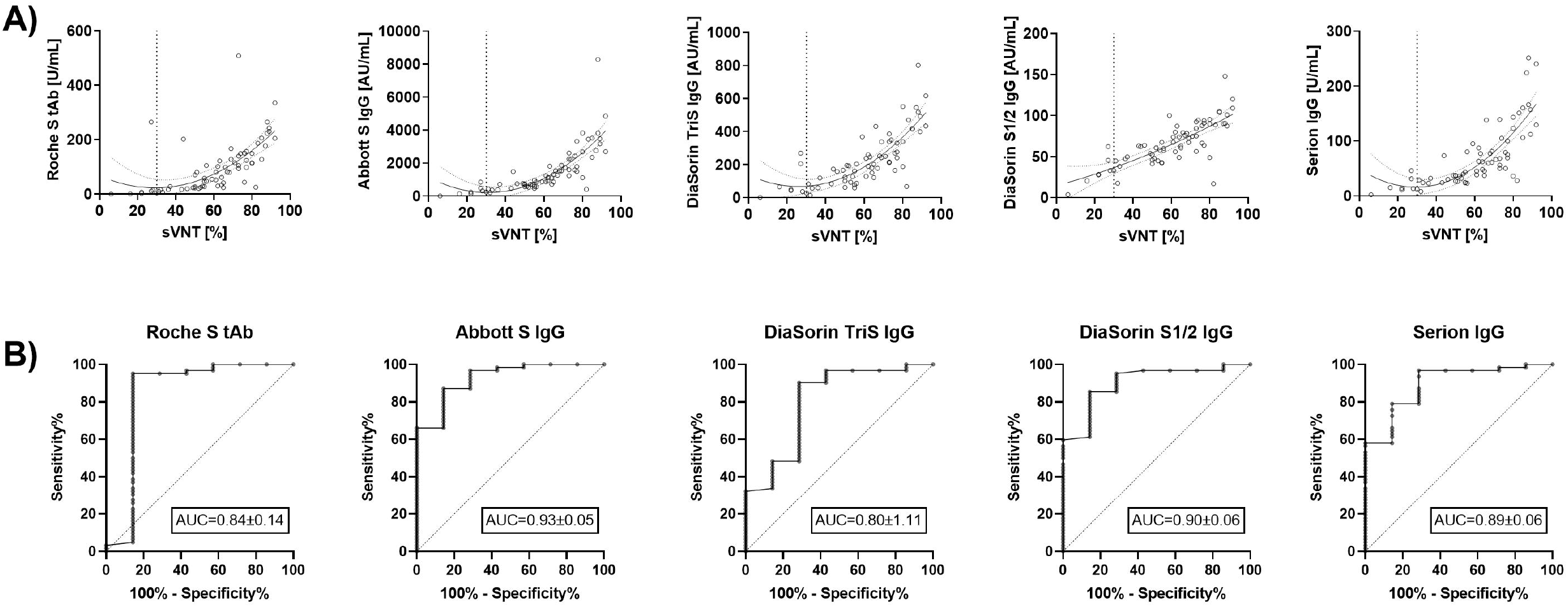
**(A)** Comparison of binding assay results converted to BAU/mL (binding antibody units per milliliter). Given are linear regression curves and their 95% confidence intervals. Dotted diagonal lines represent lines of equality. **(B)** ROC (receiver-operating-characteristics) curves of binding assays regarding the agreement with the results of a surrogate virus neutralization test (threshold for positivity: 30% inhibition).

7 (10%) of the individuals yielded sVNT results below 30% inhibition, which is considered negative according to the manufacturer (Figure 1). Binding assay results were compared between positives and negatives in the sVNT by ROC-curve analysis. The resulting AUCs ranged between 0.84 and 0.93 (see Figure 4b), however, the differences between AUCs were not statistically significant. Optimal cut-offs according to Youden’s Index, as well as other cut-offs observed from the data and their respective sensitivities and specificities are given in Supplemental Table 1. For three out of the five assays (Abbott S IgG, DiaSorin S1/2 IgG, and Serion IgG), all samples with a negative sVNT result could be correctly identified with a corresponding sensitivity of approximately 60%.

## Discussion

SARS-CoV-2 antibody assays become important tools to evaluate the proportion of people affected by COVID-19 and identify those who are still at infection risk. Now with the first vaccines available a new field of use for SARS-CoV-2 antibody tests will open up. First, many vaccinated individuals will be interested in confirming their own vaccination success based on the detection of specific antibodies. Second, vaccination-induced antibodies may be used as surrogate from which a protection correlate will be estimated. To date, only limited information on the performance of quantitative SARS-CoV-2 antibody assays is available, since most currently evaluated assays were developed in-house, as recently summarized by the CDC COVID-19 response group *(32)*. Only for a few commercially available quantitative CE-marked test systems preliminary data on the performance are given in the literature *(19, 22, 33-35)*.

Although a protection correlate for immunity in SARS-CoV-2 has not been defined yet, it is useful to begin this important preliminary work now *(6)*. Therefore, in the present work, we compared different commercial SARS-CoV-2 antibody assays with spike protein reactivity using a vaccination cohort to give a first insight into the comparability of these assays.

With regard to the numerical results, we were able to determine a broad distribution of the result values for each individual test system, so that these were presented on a logarithmic scale. This is in line with recently published reports, showing the antibody response after a single dose of BNT162b2 vaccine *(3, 4)*. Interestingly, in agreement with a study involving >500 participants in an identical study setting, we observed very similar mean values for the measurements with the DiaSorin S1/S2 IgG: 66.3 AU/mL versus 68.6 AU/mL *(3)*. Therefore, it is reasonable to assume that our cohort is representative despite the moderate number of participants. In addition, we were able to show that the results of the different test systems varied by a factor of up to more than 50. This leads to the initial conclusion that a direct comparability of the numerical results of different test systems is unlikely to be given across the range of individual findings. Differences also occurred with respect to measurement ranges, and upper measurement limits were exceeded in 2 out of 5 systems (DiaSorin TriS IgG and Serion IgG), although the study cohort reflects the antibody response before the administration of the 2^nd^ dose of Pfizer/BioNTech vaccine in SARS-CoV-2 naïve individuals. However, it must be mentioned that it is not yet known up to which level a differentiation of the obtained values is meaningful. A recently published paper shows that an anti-S post-vaccination titer of 61.8 AU/mL for the Abbott S IgG was associated with reinfection after a single dose of vaccination. Since this value is only just above the threshold for positivity for this specific assay (50 AU/mL) and in our cohort >95% of all observed values were far higher (5th percentile: 207.5 AU/mL), this finding is plausible and suggests vaccine failure due to very low antibody production. Alternatively, a reinfection might have been caused in these subjects by a virus variant where vaccination protection is mitigated. Nevertheless, it can be assumed that the average values of completely vaccinated persons are significantly higher than those in our collective and thus the upper measurement limits could frequently be exceeded in most assays. If clinically relevant, this could make additional dilution steps necessary, which are not yet taken into account by the manufacturers.

Despite the different levels of measurement, all systems showed good correlations with each other. When the measured values of the individual antibody tests were assigned to tertiles, good agreement was shown between the lowest third, the middle third and the highest third of the results. Thus, one individual with known immunosuppressive therapy consistently showed no formation of antibodies in all five antibody binding assays tested. With defined cut-offs for low or high vaccination titers of the different test systems, at least a partial transferability of a result from one to another test system may therefore be expected.

Such transferability of results could also be anticipated via referencing the antibody assays used to an international reference standard *(32)*. Indeed, a first WHO international SARS-CoV-2 antibody standard with the valence of 1000 BAU/mL has recently become available. This standard was used by the manufacturers for four out of five of the assays studied. However, this standardization was not introduced during the establishment of the test system, but post-hoc as a reference material to define a conversion factor of their own units in BAU/mL. It is therefore not surprising that, this subsequent correction did not reduce the existing systematic deviations (Figure 2) between the different tests. Only the Roche S tAb and Serion IgG tests were able to approximate the equivalence line, although here a very wide scattering of values around the trend lines was observed.

The in vitro binding of infection-associated antibodies to pathogen-specific antigens in an antibody test are important markers to objectify a past infection or vaccination. However, these do not necessarily say anything about the function of these antibodies *(1)*. In SARS-CoV-2 vaccination, an important goal is to induce neutralizing antibodies that will prevent the virus from binding to the cellular receptor, the ACE2 receptor, via the surface spike protein *(36, 37)*. Tests to neutralize live viruses can only be performed in very specialized laboratories and unfortunately, in the case of SARS-CoV-2, are not standardized, making comparability almost impossible. For this reason, we chose to use a well-characterized surrogate virus neutralization test (sVNT) as a functional reference *(38-40)*. In this assay, a simple ELISA format is used to determine the inhibition of conjugated RBD protein by neutralizing antibodies to the plate-bound ACE2 receptor. The manufacturer suggests a threshold for positivity of 30% inhibition. For the Abbott S IgG, the DiaSorin S1/2 IgG, and the Serion IgG assay, all samples below this threshold were identified at a corresponding sensitivity of about 60%. The corresponding criteria were 3 – 17 times higher than the respective assays’ thresholds for positivity. This implies that the cut-off values given for the respective test systems are only valid for the diagnosis of a past infection, but do not necessarily represent a threshold value for the presence of sufficient neutralizing activity. For the Roche S tAb and the DiaSorin TriS IgG assays, a single outlier avoided reaching maximum specificity before sensitivity dropped to 3 and 32%, respectively.

In conclusion, we found a good correlation between all evaluated assays, however, the values from the different test systems were not interchangeable, even when converted to BAU/mL using the WHO international standard for SARS-CoV-2 immunoglobulin. Furthermore, it should be noted that the thresholds for positivity provided by the manufacturers are of diagnostic value and are not indicative of sufficient inhibitory capacities.

## Data Availability

Data is available to interested researchers upon request from the corresponding author.

## Acknowledgments

We thank all sample donors for their valuable contribution. We further want to thank Miss Manuela Repl, Miss Susanne Keim, Miss Martina Trella, Miss Borka Radovanovic-Petrova, Miss Monika Martiny, Miss Jadwiga Konarski, Mr. Bernhard Haunold, Miss Maedeh Iravany, and Miss Shohreh Lashgari for perfect technical and administrative assistance. Conflicts of Interest: NP received a travel grant from DiaSorin. The Dept. of Laboratory Medicine received compensations for advertisement on scientific symposia from Roche, DiaSorin and Abbott, and holds a grant for evaluating an in-vitro diagnostic device from Roche. The GenScript sVNT test kit and the Serion IgG kit were kindly provided by the respective supplier (medac GmbH and DiaChrom), the Abbott S IgG kit and the DiaSorin TriS IgG kit were kindly provided by the manufacturers. There was no additional funding received for the present work.

## Abbreviations

SARS-CoV-2: Severe acute respiratory syndrome Coronavirus 2
COVID-19: Coronavirus Disease 19
NC: Nucleocapsid
S: Spike protein
tAb: total antibody
ECLIA: electrochemiluminescence immunoassay
RBD: receptor binding domain
CMIA: chemiluminescence microparticle assay
CLIA: chemiluminescence immunoassay
ELISA: enzyme-linked immunosorbent assay
BAU/Ml: binding antibody units per milliliter
sVNT: surrogate virus neutralization test
ROC-AUC: receiver-operating-charateristics area under the curve

## References

1. Krammer F, Simon V. Serology assays to manage covid-19. Science 2020;368:1060–1.

2. Alter G, Seder R. The power of antibody-based surveillance. N Engl J Med 2020;383:1782–4.

3. Abu Jabal K, Ben-Amram H, Beiruti K, Batheesh Y, Sussan C, Zarka S, Edelstein M. Impact of age, ethnicity, sex and prior infection status on immunogenicity following a single dose of the bnt162b2 mrna covid-19 vaccine: Real-world evidence from healthcare workers, israel, december 2020 to january 2021. Euro Surveill 2021;26.

4. Prendecki M, Clarke C, Brown J, Cox A, Gleeson S, Guckian M, et al. Effect of previous sars-cov-2 infection on humoral and t-cell responses to single-dose bnt162b2 vaccine. Lancet 2021.

5. Jin P, Li J, Pan H, Wu Y, Zhu F. Immunological surrogate endpoints of covid-2019 vaccines: The evidence we have versus the evidence we need. Signal Transduction and Targeted Therapy 2021;6:48.

6. McMahan K, Yu J, Mercado NB, Loos C, Tostanoski LH, Chandrashekar A, et al. Correlates of protection against sars-cov-2 in rhesus macaques. Nature 2020.

7. Perkmann T, Perkmann-Nagele N, Breyer MK, Breyer-Kohansal R, Burghuber OC, Hartl S, et al. Side-by-side comparison of three fully automated sars-cov-2 antibody assays with a focus on specificity. Clin Chem 2020;66:1405–13.

8. Pollan M, Perez-Gomez B, Pastor-Barriuso R, Oteo J, Hernan MA, Perez-Olmeda M, et al. Prevalence of sars-cov-2 in spain (ene-covid): A nationwide, population-based seroepidemiological study. Lancet 2020;396:535–44.

9. Oved K, Olmer L, Shemer-Avni Y, Wolf T, Supino-Rosin L, Prajgrod G, et al. Multi-center nationwide comparison of seven serology assays reveals a sars-cov-2 non-responding seronegative subpopulation. EClinicalMedicine 2020;29:100651.

10. Long QX, Tang XJ, Shi QL, Li Q, Deng HJ, Yuan J, et al. Clinical and immunological assessment of asymptomatic sars-cov-2 infections. Nat Med 2020;26:1200–4.

11. Lumley SF, Wei J, O’Donnell D, Stoesser NE, Matthews PC, Howarth A, et al. The duration, dynamics and determinants of sars-cov-2 antibody responses in individual healthcare workers. Clin Infect Dis 2021.

12. Muecksch F, Wise H, Batchelor B, Squires M, Semple E, Richardson C, et al. Longitudinal analysis of serology and neutralizing antibody levels in covid19 convalescents. J Infect Dis 2020.

13. Robbiani DF, Gaebler C, Muecksch F, Lorenzi JCC, Wang Z, Cho A, et al. Convergent antibody responses to sars-cov-2 in convalescent individuals. Nature 2020;584:437–42.

14. Buss LF, Prete CA Jr.,, Abrahim CMM, Mendrone A Jr.,, Salomon T, de Almeida-Neto C, et al. Three-quarters attack rate of sars-cov-2 in the brazilian amazon during a largely unmitigated epidemic. Science 2021;371:288–92.

15. Bolotin S, Tran V, Osman S, Brown KA, Buchan SA, Joh E, et al. Sars-cov-2 seroprevalence survey estimates are affected by anti-nucleocapsid antibody decline. J Infect Dis 2021.

16. Favresse J, Brauner J, Bodart N, Vigneron A, Roisin S, Melchionda S, et al. An original multiplex method to assess five different sars-cov-2 antibodies. Clin Chem Lab Med 2020.

17. Le Bert N, Tan AT, Kunasegaran K, Tham CYL, Hafezi M, Chia A, et al. Sars-cov-2-specific t cell immunity in cases of covid-19 and sars, and uninfected controls. Nature 2020;584:457–62.

18. Musico A, Frigerio R, Mussida A, Barzon L, Sinigaglia A, Riccetti S, et al. Sars-cov-2 epitope mapping on microarrays highlights strong immune-response to n protein region. Vaccines (Basel) 2021;9.

19. Klausberger M, Dürkop M, Haslacher H, Wozniak-Knopp G, Cserjan-Puschmann M, Perkmann T, et al. A comprehensive antigen production and characterization study for easy-to-implement, highly specific and quantitative sars-cov-2 antibody assays. medRxiv 2021.

20. Wajnberg A, Amanat F, Firpo A, Altman DR, Bailey MJ, Mansour M, et al. Robust neutralizing antibodies to sars-cov-2 infection persist for months. Science 2020;370:1227–30.

21. Bal A, Pozzetto B, Trabaud MA, Escuret V, Rabilloud M, Langlois-Jacques C, et al. Evaluation of high-throughput sars-cov-2 serological assays in a longitudinal cohort of patients with mild covid-19: Clinical sensitivity, specificity and association with virus neutralization test. Clin Chem 2021.

22. Bonelli F, Sarasini A, Zierold C, Calleri M, Bonetti A, Vismara C, et al. Clinical and analytical performance of an automated serological test that identifies s1/s2-neutralizing igg in covid-19 patients semiquantitatively. J Clin Microbiol 2020;58.

23. Padoan A, Bonfante F, Pagliari M, Bortolami A, Negrini D, Zuin S, et al. Analytical and clinical performances of five immunoassays for the detection of sars-cov-2 antibodies in comparison with neutralization activity. EBioMedicine 2020;62:103101.

24. Patel EU, Bloch EM, Clarke W, Hsieh YH, Boon D, Eby Y, et al. Comparative performance of five commercially available serologic assays to detect antibodies to sars-cov-2 and identify individuals with high neutralizing titers. J Clin Microbiol 2021;59.

25. Suhandynata RT, Hoffman MA, Huang D, Tran JT, Kelner MJ, Reed SL, et al. Commercial serology assays predict neutralization activity against sars-cov-2. Clin Chem 2020.

26. Tang MS, Case JB, Franks CE, Chen RE, Anderson NW, Henderson JP, et al. Association between sars-cov-2 neutralizing antibodies and commercial serological assays. Clin Chem 2020.

27. Krammer F. Sars-cov-2 vaccines in development. Nature 2020;586:516–27.

28. Nie J, Li Q, Wu J, Zhao C, Hao H, Liu H, et al. Establishment and validation of a pseudovirus neutralization assay for sars-cov-2. Emerg Microbes Infect 2020;9:680–6.

29. Oguntuyo KY, Stevens CS, Hung CT, Ikegame S, Acklin JA, Kowdle SS, et al. Quantifying absolute neutralization titers against sars-cov-2 by a standardized virus neutralization assay allows for cross-cohort comparisons of covid-19 sera. mBio 2021;12.

30. Vanderheiden A, Edara VV, Floyd K, Kauffman RC, Mantus G, Anderson E, et al. Development of a rapid focus reduction neutralization test assay for measuring sars-cov-2 neutralizing antibodies. Curr Protoc Immunol 2020;131:e116.

31. Haslacher H, Gerner M, Hofer P, Jurkowitsch A, Hainfellner J, Kain R, et al. Usage data and scientific impact of the prospectively established fluid bioresources at the hospital-based meduni wien biobank. Biopreserv Biobank 2018;16:477–82.

32. Gundlapalli AV, Salerno RM, Brooks JT, Averhoff F, Petersen LR, McDonald LC, et al. Sars-cov-2 serologic assay needs for the next phase of the us covid-19 pandemic response. Open Forum Infect Dis 2021;8:ofaa555.

33. Schaffner A, Risch L, Aeschbacher S, Risch C, Weber MC, Thiel SL, et al. Characterization of a pan-immunoglobulin assay quantifying antibodies directed against the receptor binding domain of the sars-cov-2 s1-subunit of the spike protein: A population-based study. J Clin Med 2020;9.

34. Soleimani R, Khourssaji M, Gruson D, Rodriguez-Villalobos H, Berghmans M, Belkhir L, et al. Clinical usefulness of fully automated chemiluminescent immunoassay for quantitative antibody measurements in covid-19 patients. J Med Virol 2021;93:1465–77.

35. Higgins V, Fabros A, Kulasingam V. Quantitative measurement of anti-sars-cov-2 antibodies: Analytical and clinical evaluation. J Clin Microbiol 2021.

36. Yang J, Petitjean SJL, Koehler M, Zhang Q, Dumitru AC, Chen W, et al. Molecular interaction and inhibition of sars-cov-2 binding to the ace2 receptor. Nat Commun 2020;11:4541.

37. Yan R, Zhang Y, Li Y, Xia L, Guo Y, Zhou Q. Structural basis for the recognition of sars-cov-2 by full-length human ace2. Science 2020;367:1444–8.

38. Abe KT, Li Z, Samson R, Samavarchi-Tehrani P, Valcourt EJ, Wood H, et al. A simple protein-based surrogate neutralization assay for sars-cov-2. JCI Insight 2020;5.

39. Perera R, Ko R, Tsang OTY, Hui DSC, Kwan MYM, Brackman CJ, et al. Evaluation of a sars-cov-2 surrogate virus neutralization test for detection of antibody in human, canine, cat, and hamster sera. J Clin Microbiol 2021;59.

40. von Rhein C, Scholz T, Henss L, Kronstein-Wiedemann R, Schwarz T, Rodionov RN, et al. Comparison of potency assays to assess sars-cov-2 neutralizing antibody capacity in covid-19 convalescent plasma. J Virol Methods 2021;288:114031.

