## Supplemental Table 1 for "Anti-Spike protein assays to determine post-vaccination antibody levels: a head-to-head comparison of five quantitative assays"

|  | Roche S tAb | | | Abbott S IgG | | | DiaSorin TriS IgG | | |
| --- | --- | --- | --- | --- | --- | --- | --- | --- | --- |
| ROC AUC | 0.84±0.14 | | | 0.93±0.05 | | | 0.80±1.11 | | |
| Youden’s Index | 0.81 | | | 0.73 | | | 0.62 | | |
| Associated Criterion | >15.0 | | | >476.1 | | | >65.2 | | |
|  |  | **Sensitivity** | **Specificity** |  | **Sensitivity** | **Specificity** |  | **Sensitivity** | **Specificity** |
|  |  | 95.2% (86.5 - 99.0) | 85.7% (42.1 - 99.6) |  | 87.1% (76.1 - 94.3) | 85.7% (42.1 - 99.6) |  | 90.3% (80.1 - 96.4) | 71.4% (29.0 - 96.3) |
|  | **Selected Criteria:** | |  | **Selected Criteria:** | |  | **Selected Criteria:** | |  |
|  | ≥0.4 | 100% (94.2 - 100.0) | 0% (0.0 - 41.0) | ≥1.4 | 100% (94.2 - 100.0) | 0% (0.0 - 41.0) | ≥1.8 | 100% (94.2 - 100.0) | 0% (0.0 - 41.0) |
|  | >2.6 | 100% (94.2 - 100.0) | 42.9% (9.9 - 81.6) | >140.0 | 100% (94.2 - 100.0) | 42.9% (9.9 - 81.6) | >1.8 | 100% (94.2 - 100.0) | 14.3% (0.4 - 57.9) |
|  | >6.6 | 96.8% (88.8 - 99.6) | 42.9% (9.9 - 81.6) | >211.0 | 98.4% (91.3 - 100.0) | 42.9% (9.9 - 81.6) | >28.4 | 96.8% (88.8 - 99.6) | 14.3% (0.4 - 57.9) |
|  | >7.2 | 96.8% (88.8 - 99.6) | 57.1% (18.4 - 90.1) | >233.8 | 98.4% (91.3 - 100.0) | 57.1% (18.4 - 90.1) | >48.8 | 96.8% (88.8 - 99.6) | 57.1% (18.4 - 90.1) |
|  | >10.1 | 95.2% (86.5 - 99.0) | 57.1% (18.4 - 90.1) | >244.8 | 96.8% (88.8 - 99.6) | 57.1% (18.4 - 90.1) | >61.9 | 90.3% (80.1 - 96.4) | 57.1% (18.4 - 90.1) |
|  | >15.0 | 95.2% (86.5 - 99.0) | 85.7% (42.1 - 99.6) | >315.6 | 96.8% (88.8 - 99.6) | 71.4% (29.0 - 96.3) | >65.2 | 90.3% (80.1 - 96.4) | 71.4% (29.0 - 96.3) |
|  | >243.0 | 4.8% (1.0 - 13.5) | 85.7% (42.1 - 99.6) | >472.4 | 87.1% (76.1 - 94.3) | 71.4% (29.0 - 96.3) | >203.0 | 48.4% (35.5 - 61.4) | 71.4% (29.0 - 96.3) |
|  | >265.0 | 3.2% (0.4 - 11.2) | 100% (59.0 - 100.0) | >476.1 | 87.1% (76.1 - 94.3) | 85.7% (42.1 - 99.6) | >205.0 | 48.4% (35.5 - 61.4) | 85.7% (42.1 - 99.6) |
|  | >508.0 | 0% (0.0 - 5.8) | 100% (59.0 - 100.0) | >763.2 | 66.1% (53.0 - 77.7) | 85.7% (42.1 - 99.6) | >259.0 | 33.9% (22.3 - 47.0) | 85.7% (42.1 - 99.6) |
|  |  |  |  | >847.1 | 66.1% (53.0 - 77.7) | 100% (59.0 - 100.0) | >269.0 | 32.3% (20.9 - 45.3) | 100% (59.0 - 100.0) |
|  |  |  |  | >8281.0 | 0% (0.0 - 5.8) | 100% (59.0 - 100.0) | >ULoQ | 0% (0.0 - 5.8) | 100% (59.0 - 100.0) |
|  | **DiaSorin S1/2 IgG** | | | **Serion IgG** | | |  |  |  |
| ROC AUC | 0.90±0.06 | | | 0.89±0.06 | | |  |  |  |
| Youden’s Index | 0.71 | | | 0.68 | | |  |  |  |
| Associated Criterion | >45.6 | | | >15 | | |  |  |  |
|  |  | **Sensitivity** | **Specificity** |  | **Sensitivity** | **Specificity** |  |  |  |
|  |  | 85.5% (74.2 - 93.1) | 85.7% (42.1 - 99.6) |  | 96.8% (88.8 - 99.6) | 71.4% (29.0 - 96.3) |  |  |  |
|  | **Selected Criteria:** | |  | **Selected Criteria:** | |  |  |  |  |
|  | ≥LLoQ | 100% (94.2 - 100.0) | 0% (0.0 - 41.0) | ≥2 | 100% (94.2 - 100.0) | 0% (0.0 - 41.0) |  |  |  |
|  | >LLoQ | 100% (94.2 - 100.0) | 14.3% (0.4 - 57.9) | >2 | 100% (94.2 - 100.0) | 14.3% (0.4 - 57.9) |  |  |  |
|  | >17.6 | 96.8% (88.8 - 99.6) | 14.3% (0.4 - 57.9) | >8 | 98.4% (91.3 - 100.0) | 14.3% (0.4 - 57.9) |  |  |  |
|  | >28.4 | 96.8% (88.8 - 99.6) | 57.1% (18.4 - 90.1) | >11 | 98.4% (91.3 - 100.0) | 28.6% (3.7 - 71.0) |  |  |  |
|  | >33.0 | 95.2% (86.5 - 99.0) | 71.4% (29.0 - 96.3) | >12 | 96.8% (88.8 - 99.6) | 28.6% (3.7 - 71.0) |  |  |  |
|  | >44.9 | 85.5% (74.2 - 93.1) | 71.4% (29.0 - 96.3) | >15 | 96.8% (88.8 - 99.6) | 71.4% (29.0 - 96.3) |  |  |  |
|  | >45.6 | 85.5% (74.2 - 93.1) | 85.7% (42.1 - 99.6) | >30 | 79% (66.8 - 88.3) | 71.4% (29.0 - 96.3) |  |  |  |
|  | >60.8 | 61.3% (48.1 - 73.4) | 85.7% (42.1 - 99.6) | >31 | 79% (66.8 - 88.3) | 85.7% (42.1 - 99.6) |  |  |  |
|  | >62.2 | 59.7% (46.4 - 71.9) | 100% (59.0 - 100.0) | >45 | 58.1% (44.8 - 70.5) | 85.7% (42.1 - 99.6) |  |  |  |
|  | >148 | 0% (0.0 - 5.8) | 100% (59.0 - 100.0) | >46 | 58.1% (44.8 - 70.5) | 100% (59.0 - 100.0) |  |  |  |
|  |  |  |  | >ULoQ | 0% (0.0 - 5.8) | 100% (59.0 - 100.0) |  |  |  |

**Supplemental Table 1:** Results from receiver-operating-characteristics (ROC)-analysis for five different binding assays. AUCs (areas under the curves), Youden’s Index and sensitivities/specificities for the criterion associated with the Youden’s Index (in U/mL or AU/mL, respectively), as well as for selected criterion values are given. LLoQ… Lower limit of quantification, ULoQ… Upper limit of quantification.
